# New Insight into the Neural Mechanisms of Migraine in Adolescents: Relationships with Sleep

**DOI:** 10.1101/2021.04.09.21255214

**Authors:** Hadas Nahman-Averbuch, Victor J Schneider, Gregory R. Lee, James L. Peugh, Andrew D. Hershey, Scott W. Powers, Massimiliano de Zambotti, Robert C. Coghill, Christopher D. King

## Abstract

Adolescents with migraine have different functional connectivity of the amygdala compared to individuals without migraine. Considering that sleep is often disturbed in those adolescents with migrane, this study examined if measures of subjective and objective (actigraphic) sleep difficulties mediate alterations in amygdalar connectivity in adolescents with migraine compared to healthy adolescents. Twenty adolescents with migraine and 20 healthy controls completed surveys about their headaches and overall sleep quality, sleep hygiene and perceived sleep difficulties, wore a wrist-worn actigraphy, and underwent an MRI scan.

Adolescents with migraine differed from healthy controls only in perceived sleep difficulties related to sleep initiation and maintenance (*p* <0.01) and had greater functional connectivity between the amygdala and the posterior cingulate cortex, precuneus, dorsolateral prefrontal, sensorimotor, and the occipital cortexes. While the mediation model showed group differences in subjective and actigraphic sleep difficulties, these did not mediate the differences in amygdalar connectivity found between the groups. Adolescents with migraine have greater connectivity between the amygdala and areas involved in sensory, affective, and cognitive aspects of pain. These alterations may not be due to higher levels of sleep difficulties in adolescents with migraine, suggesting that both amygdala and sleep alterations may play an independent role in migraine pathophysiology

**Perspective:** This article evaluates the role plays by sleep on neural alterations in adolescents with migraine. It indicates that neural alterations due to migraine are not related to alterations in subjective and actigraphic sleep difficulties. This advances the understanding of the mechanisms underlying pediatric migraine and can potentially advance migraine management.

## Introduction

Migraine is one of the most prevalent and disabling pain conditions (1, 2). Various studies have found alterations in brain structure and function in participants with migraine compared to healthy controls (for review (3-6)). Only a few studies have examined neural alterations in youth with migraine (7-9). These studies found alterations in the function of brain regions such as the amygdala, insula, primary somatosensory cortex (SI) and thalamus, but were somewhat limited by a wide age range (including young adults) as well as by having participants on preventive medications. Thus, the neural mechanisms involved in migraine in adolescents remain to be fully characterized.

One factor, which has frequently been associated with elevated pain in healthy adolescents and their peers with pain, is sleep (10-13). Clinically, individuals with chronic pain often report disrupted sleep (e.g., poor sleep quality, shorter sleep duration, insomnia symptoms) (14-19), which can be accompanied by increases in the severity of clinical pain and sensitivity to experimental pain (17, 20). In adults with migraine, the literature has focused on naturally occurring conditions related to sleep deprivation (e.g., not enough time spent asleep) and continuity disturbances (e.g., irregular sleep patterns, increased nightly awakenings) as precipitating factors for elicitation of migraine (21, 22). Similar findings have been observed with pediatric patients with migraine as disturbed sleep (e.g., short sleep durations, poorer sleep quality) is often associated with greater headache frequency and disability (23, 24). The extent to which, and the neurobiological mechanisms by which sleep disruption exacerbates pain in people with migraine remain almost wholly unexplored. One study examined this in adult participants with migraine and found that amygdalar connectivity was related to the severity of sleep disturbances (measured as categorical four-grade sleep disturbance scale ranging from normal (0) to serious sleep disturbance (3)) (25). The effect of sleep on neural function in adolescents with migraine as well is still unclear.

The aims of the present study were to (1) evaluate subjective and objective (actigraphic) sleep in adolescents with and without migraine, (2) evaluate resting-state functional connectivity of the amygdala in adolescents with and without migraine, (3) explore if measures of subjective and objective sleep mediate the differences in amygdalar connectivity in adolescents with and without migraine.

## Materials and Methods

This study is independent and not related to previous published studies involving adolescents with migraine from our group (26-28).

### Participants

Twenty adolescents with migraine (17 Females, 3 males; mean age: 14.7±1.8 yo) and twenty healthy adolescents (17 Females, 3 males; mean age: 14.8±1.6 yo) participated in the study. Participants were recruited from the Cincinnati Children’s Headache Center.

Inclusion criteria for youth with migraine: 1) Male or female patients; 2) Age 12-17 years old; 3) diagnosis of migraine with aura, migraine without aura, and/or chronic migraine that meets the International Classification of Headache Disorders, 3^rd^ Edition (ICHD-3) criteria (29) as determined by a headache medicine neurologist; 4) No other diagnosed chronic pain conditions; 5) Not taking any current migraine preventive medication (e.g., amitriptyline, topiramate, depakote); 6) English-speaking. Healthy controls were age and sex matched to patients with migraine. Inclusion criteria for healthy controls: 1) Male or female participants 2) Age 12-17 years old; 3) No diagnosed chronic pain conditions including migraine or other headache disorders.

Participants were ineligible for participation if they met the following exclusion criteria: 1) Pregnant (via urine pregnancy test) ; 2) Weight less than 30 kg (66 lbs) or greater than 120 kg (264 lbs.) or weight/size incompatible with MRI scanner; 3) Orthodontic braces or other metallic implants, which obscure or interfere with the MRI; 4) Claustrophobia; 5) Diagnosis of psychiatric conditions including depression, ADD/ADHD, and anxiety; 6) Diagnosis of neurological (e.g., epilepsy) conditions other than migraine; 7) Documented developmental delays or impairments (e.g., autism, cerebral palsy, or mental retardation); 8) Current use of prescribed medications or products including opioids, antipsychotics, antimanics, barbiturates, benzodiazepines, muscle relaxants, sedatives, tramadol, Selective Serotonin Reuptake Inhibitor (SSRIs), or Serotonin and norepinephrine reuptake inhibitors (SSNRIs).

### Study design overview

The study was approved by the Cincinnati Children’s Hospital institutional Review Board (IRB). Data were collected from October 2016 to October 2017. Informed parental consent and participant assent were obtained before study procedures began. The study included two in-lab study visits that were approximately 7-days apart. In the first study visit, participants completed surveys about their headaches and self-report measures of sleep. Participants were also given an Actiwatch 2 (Phillips Respironics) that they wore for at least 7 days. After 7 days, they returned for the second visit, which included an MRI scan. All scans were resting state scans. Scans were completed between 7:30-16:30 with no significant differences between the groups on the time of scans (p=0.756). Participants were directed to close their eyes during all scans. At the end of each scan participants were asked to rate their current headache pain intensity and unpleasantness.

Data from this study will be shared at the request of other investigators for purposes of replicating procedures and results.

### Self-reported Outcomes

All participants completed a series of questionnaires at the first study visit, including a series of questionnaires to characterize sleep difficulties (insomnia symptoms) and typical sleep habits.

#### Migraine and Health History

Participants completed a self-reported survey measuring the frequency (days with mild, moderate, or severe headaches over the past month) and the average intensity (numerical rating scale, 0-10) of migraine.

#### Functional Disability Inventory (FDI)

The FDI assesses children’s and adolescent’s self-reported difficulty in physical and psychosocial functioning due to their physical health (30). The FDI is a widely used, reliable and validated tool to assess functional disability. It contains 15 items related to activity limitations that are rated on a 5-point scale (0-no trouble; 4-impossible). Scores range from 0 to 60, with higher scores indicating greater disability. Internal consistency of FDI is _α_ = .96.

#### Pediatric Migraine Disability Assessment Scale (PedMIDAS)

This questionnaire has six items that assess the number of days headaches affected the participant’s daily activities. The days indicated by the participant are summed for a total score. This is a widely used, reliable and validated tool to assess migraine disability. Higher scores indicate higher migraine disability with scores of 0-10 indicates little to no disability, 11-30 mild disability, 31-50 moderate disability and >50 severe disability (31, 32). Internal consistency of PedMIDAS is _α_ = .83. PedMIDAS was not completed by controls as it is specific to impact of having migraine.

#### The Patient-Reported Outcomes Measurement Information System (PROMIS)

Two pediatric PROMIS surveys were used to assess emotional distress (33, 34). The PROMIS Anxiety survey included 8 items assessing fear, anxious misery, and hyperarousal. Internal consistency of PROMIS Anxiety is _α_ = .89. The PROMIS Depression survey included 8 items assessing depressive symptoms such as negative mood, anhedonia, negative views of self, and negative social cognition. Internal consistency of PROMIS Anxiety is _α_ = .79. For the current study, raw scores were compared between groups.

#### Pain intensity and unpleasantness

Before the MRI scans, participants received instructions on how to rate pain intensity and unpleasantness. Pain intensity (how strong the pain feels) and pain unpleasantness (how disturbing the pain is) were defined using a radio analogy (35). After each scan, participants were asked to indicate the magnitude of pain sensation along the visual analog scale (VAS). The VAS is a simple rating scale with ratio scale properties (36).

Participants were instructed to move a 15cm hand held slider. The further the slider was moved, the greater the perceived pain-sensation intensity / unpleasantness. Both intensity and unpleasantness dimensions ranged between no pain sensation / not unpleasant, which corresponds to a rating of 0, and the most intense pain sensation imaginable / most unpleasant imaginable, which corresponds to a rating of 10. The ratings were recorded with a precision of 0.1 units.

#### Insomnia Severity Index (ISI)

The ISI is a 7-item measure that assesses the presence, severity and impact of insomnia symptoms (e.g., difficulty failing and maintaining sleep) over the past 2 weeks (37-39). The ISI assesses sleep domains related to problems with sleep onset, sleep maintenance, daytime sleepiness, daytime impairment, and distress. Scores range from 0 to 28, with higher scores indicative of more severe sleep difficulties (≤7, no clinically significant insomnia; 8-14 subthreshold insomnia, and ≥15, clinical insomnia) (40). Internal consistency of ISI is α = .81. The ISI has been validated in both adults (41) (including headache (42)) and healthy adolescents (41), and used in pediatric pain conditions (37).

#### Adolescent Sleep Hygiene Scale (ASHS)

The ASHS is a 33-item measure assessing the frequency of sleep-promoting and sleep-inhibiting behaviors over the past month (43). The ASHS assesses the behaviors across several domains including physiological, cognitive, emotional, sleep environment, daytime sleepiness, use of substances, bedtime routine, and sleep stability. The ASHS has been used to assess sleep/wake behavioral patterns in healthy adolescents and adolescents with chronic pain conditions (44, 45). A total score was computed by averaging all items on the scale, with higher scores reflecting better sleep hygiene. Internal consistency for the full scale is α = .85.

### Objective sleep evaluation: Actigraphy

Standard actigraphy was used to characterize the objective sleep-wake patterns. Participants wore an Actiwatch 2 (Respironics, Bend, OR; http://www.actigraphy.respironics.com), providing accepted objecitve measure of sleep in non-laboratory settings, on their nondominant wrist for up to 7 days. Actigraphy data was available for all participants (M= 6.9±0.3 days, range = 5-7 days). Actiware software (version 6.0) was used to score the actigraphy data using a proprietary algorithm that scores the amplitude and frequency of detected movements every 30 seconds. At Visit 2 (MRI), the actigraphy file and sleep diary were reviewed with the participant by experienced trained lab staff and remaining discrepancies were further discussed. Actigraphy sleep onset (first time patient fell asleep for the night) and offset (time patient woke up in the morning without falling back asleep) were adjusted based on participant’s sleep diary (46, 47). If the automatic rest period determined by the Actigraphy algorithm occurred within 30 minutes of the sleep and wake times reported in the diary, the rest period was left as-is. If either the sleep or wake time occurred more than 30 minutes beyond the sleep or wake time reported in the diary, the actiwatch’s reported activity levels and the participant’s reported sleep and wake times were used to determine the most accurate interval possible. The following sleep variables were calculated and averaged across the 7-nights recording period: (a) *total sleep time* (TST; total amount of time scored as sleep from sleep onset to offset, in minutes), (b) *wake minutes after sleep onset* (WASO; time scored as wake after sleep onset, in minutes), and (c) *sleep efficiency* (SE; calculated ratio between total time sleep vs. total time spent in bed as a percentage; higher % = More efficient sleep).

### Imaging acquisition

Participants were scanned on a 3T Philips Ingenia scanner with a 32 Channel head coil. The total scan time was one hour. One T1 anatomical scan, two resting state pseudo-continuous arterial spin labeling (pCASL) and two resting state BOLD scans were acquired. The T1 anatomical scan was always completed first. Then the two BOLD or pCASL scans were completed in a random order based on the participant number. Details on the pCASL analysis and results are summarized in Supplementary 1. A radiologist reviewed the MRI scans in order to detect incidental findings. No major incidental findings were found.

High-resolution T1: multiecho (4 echoes) weighted images were obtained using the following parameters: TR=10ms, TE=1.8, 3.8, 5.8, 7.8ms, field of view was 256×224×200mm, voxel size=1×1×1mm, number of slices=200, flip angle=8 degrees, slice orientation=sagittal, total scan time=4:42 minutes.

BOLD fMRI was performed to examine resting state connectivity using FOV=240×240mm, voxel size=3×3mm, slice thickness=4mm, 34 slices, flip angle= 90 degrees, TE=35ms, TR= 2000ms, number of volumes=193, slice orientation=transverse, slice order=ascending, dummy scans=2 (data not saved during export from scanner), total scan duration=6:34 minutes.

### Image processing and statistical analysis

The FSL (FMRIB’s Software Library, Oxford, UK, version 5.0.8) software package was used for the vast majority of image processing operations and statistical analyses and was augmented by scripts.

T1-weighted images were first bias corrected using FMRIB’s Automated Segmentation Tool, FAST. The T1-weighted images were brain extracted using the brain extraction tool (BET) and normalized into standard anatomic space (MNI152) using FLIRT (FMRIB’s Linear Image Registration Tool) (48). Then, images were segmented into the different tissue types and masked with a probability threshold of 0.95 for white matter and CSF.

#### BOLD / Connectivity Analyses

For EPI images, we first created a mask of the brain. We then employed slice timing correction using slicetimer and outlier detection using FSL motion outliers routines. Corrupted volumes were identified, and these time points were regressed out of the analysis. In this data set, the mean corrupted volumes per participant were 7.6±4.4 and 8.4±4.3 in adolescents with migraine vs. healthy controls, respectively (p=0.566). Absolute mean head motion across the BOLD series was not different between the groups (0.41±0.27 mm vs. 0.39±0.21 mm in the migraine and healthy groups respectvely, p=0.0744).

Images were then co-registered to the high resolution T1 structural image and normalized using FMRIB’s Linear Image Registration Tool (FLIRT). A component-based noise correction method (aCompCor) was used to minimize the impact of physiological, scanner and motion-related artifacts (49). A principal component analysis (PCA) was used to characterize the time series data from the white matter and CSF. The top 5 principal components were then introduced as covariates in a general linear model (GLM). In addition, 24 motion regressors were included in the GLM design. These regressors were calculated using mcFLIRT and included three regressors for rotation and three for translation. These six regressors were: 1) 1 volume delayed, 2) demeaned and squared, 3) delayed multiplied by the demeaned squared to give a total of 24 motion-related regressors. Denoised residuals were then imported into FEAT and were smoothed with a 5 mm filter and underwent highpass temporal filtering (>0.01 Hz). These images then underwent intensity normalization of each volume in the series. FLIRT was used to register functional images to the high resolution structural image using boundary based registration (50), and to the standard space (MNI152) with warp resolution of 10mm. Registration from high resolution space to standard space was performed using FNIRT (51). BOLD images were then placed in standard space by combining the BOLD to structural and the structural to standard transformations.

Time courses of activity were extracted from seed regions. Regions of the right and left amygdala were chosen as seeds based on previous literature suggesting involvement of the amygdala in migraine pathophysiology (25, 52) and in behavioral therapy in adolescents (53) The Juelich atlas was used to create a mask of the seed regions of the right and left amygdala using 50% probability.

The time courses of each seed were separately analyzed using FILM (FMRIB’s Improved Linear Model). First level, fixed-effects analyses were run for each BOLD series to identify voxels that had time courses that were significantly positively and negatively correlated with that of the amygdala seed (i.e., seed to whole brain analysis). Second level fixed effects analyses were used to examine within participants effects across imaging series. Finally, third level random effects analyses (FMRIB’s Local Analysis of Mixed Effects, FLAME 1+2) were employed to compare differences in functional connectivity between adolescents with migraine and healthy controls. Clusters of connectivity were identified using a threshold of Z>2.3 and corrected cluster significance of p<0.05 were estimated according to Gaussian random field theory (54).

Mediation models were used to examine if measures of perceived sleep difficulties and objective sleep disruption mediate the differences in amygdalar connectivity between the groups. In order to avoid type 1 error inflation (by separate examination of the several multiple sleep parameters assessed), we included ISI, reflecting subjective measure for sleep difficulties and significantly differing betwewn groups, and actigraphic WASO, reflecting objective sleep disruption. Separate mediation analyses were conducted using bootstrapping with ISI/WASO scores as the mediator. Featquery was used to extract the parameter estimate for each participant using a mask that included the significant differences in functional connectivity between the groups (separately for the left and right amygdala).

For demographic and behavioral data, T-tests with FDR Type-1 error control were used to compare demographic, pain, actigraphy and perceived sleep measures and psychological data between adolescents with migraine and healthy controls using SPSS (version 24). Equality of variances was tested using the Levene’s Test.

## Results

### Pain characteristics in adolescents with and without migraine

Participant’s demographic, pain, sleep and psychological data are summarized in Table 1. Adolescents with migraine had greater disability based on FDI (p=0.001, Fig. 1A), and greater pain intensity ratings (p=0.009, Fig. 1B) and pain unpleasantness ratings (p=0.013, Fig. 1C) at the beginning of the scan compared to healthy adolescents. It is of worth to note that althought the differences in pain ratings were significantly different between the groups, they were mild. No differences between the groups were found for anxiety and depression levels (*p*’s < 0.10). Three adolescents with migraine took over the counter pain medications within 24 hours before the scan (ibuprofen, naproxen).

**Table 1.**
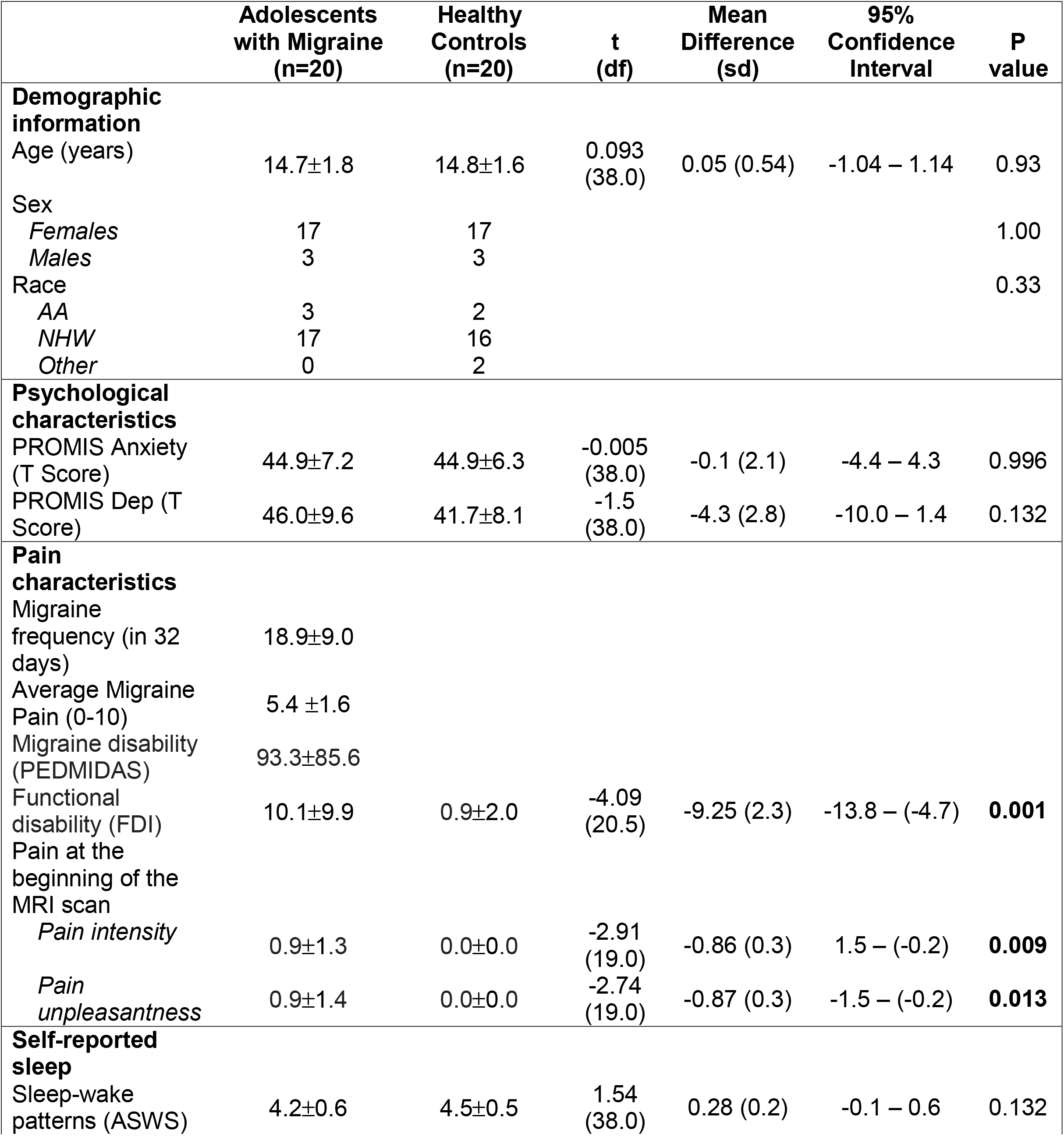
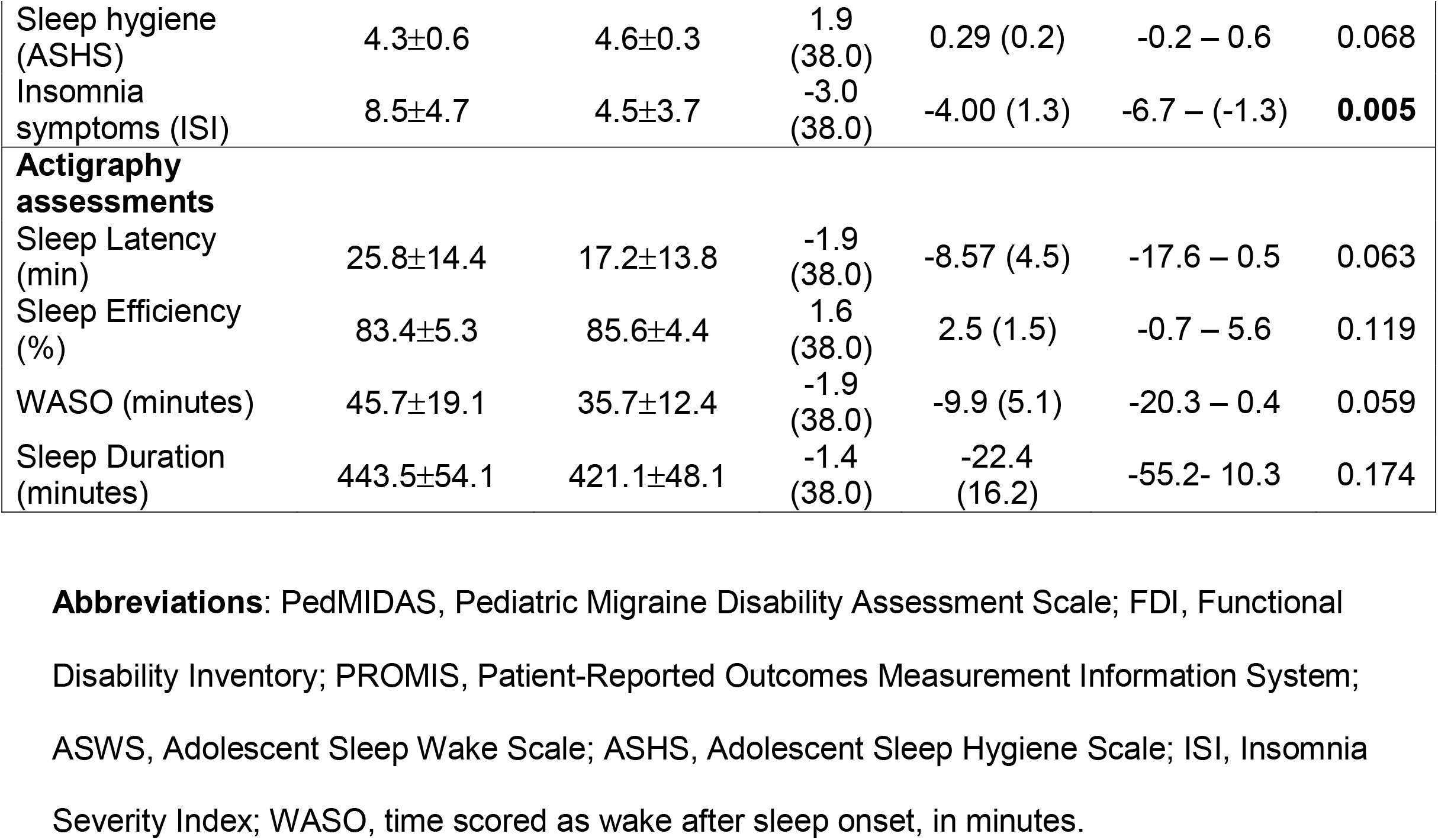
Differences between adolescents with and without migraine.

|  | **Adolescents**<br>**with Migraine**<br>**(n=20)** | **Healthy**<br>**Controls**<br>**(n=20)** | **t**<br>**(df)** | **Mean**<br>**Difference**<br>**(sd)** | **95%**<br>**Confidence**<br>**Interval** | **P**<br>**value** |
| --- | --- | --- | --- | --- | --- | --- |
| **Demographic**<br>**information** |  |  |  |  |  |  |
| Age (years) | 14.7±1.8 | 14.8±1.6 | 0.093<br>(38.0) | 0.05 (0.54) | -1.04 – 1.14 | 0.93 |
| Sex |  |  |  |  |  |  |
| *Females* | 17 | 17 |  |  |  | 1.00 |
| *Males* | 3 | 3 |  |  |  |  |
| Race |  |  |  |  |  | 0.33 |
| *AA* | 3 | 2 |  |  |  |  |
| *NHW* | 17 | 16 |  |  |  |  |
| *Other* | 0 | 2 |  |  |  |  |
| **Psychological**<br>**characteristics** |  |  |  |  |  |  |
| PROMIS Anxiety<br>(T Score) | 44.9±7.2 | 44.9±6.3 | -0.005<br>(38.0) | -0.1 (2.1) | -4.4 – 4.3 | 0.996 |
| PROMIS Dep (T<br>Score) | 46.0±9.6 | 41.7±8.1 | -1.5<br>(38.0) | -4.3 (2.8) | -10.0 – 1.4 | 0.132 |
| **Pain**<br>**characteristics** |  |  |  |  |  |  |
| Migraine<br>frequency (in 32<br>days) | 18.9±9.0 |  |  |  |  |  |
| Average Migraine<br>Pain (0-10) | 5.4 ±1.6 |  |  |  |  |  |
| Migraine disability<br>(PEDMIDAS) | 93.3±85.6 |  |  |  |  |  |
| Functional<br>disability (FDI) | 10.1±9.9 | 0.9±2.0 | -4.09<br>(20.5) | -9.25 (2.3) | -13.8 – (-4.7) | **0.001** |
| Pain at the<br>beginning of the<br>MRI scan |  |  |  |  |  |  |
| *Pain intensity* | 0.9±1.3 | 0.0±0.0 | -2.91<br>(19.0) | -0.86 (0.3) | 1.5 – (-0.2) | **0.009** |
| *Pain*<br>*unpleasantness* | 0.9±1.4 | 0.0±0.0 | -2.74<br>(19.0) | -0.87 (0.3) | -1.5 – (-0.2) | **0.013** |
| **Self-reported**<br>**sleep** |  |  |  |  |  |  |
| Sleep-wake<br>patterns (ASWS) | 4.2±0.6 | 4.5±0.5 | 1.54<br>(38.0) | 0.28 (0.2) | -0.1 – 0.6 | 0.132 |
| Sleep hygiene (ASHS) | 4.3±0.6 | 4.6±0.3 | 1.9 (38.0) | 0.29 (0.2) | -0.2 – 0.6 | 0.068 |
| Insomnia symptoms (ISI) | 8.5±4.7 | 4.5±3.7 | -3.0 (38.0) | -4.00 (1.3) | -6.7 – (-1.3) | **0.005** |
| **Actigraphy assessments** |  |  |  |  |  |  |
| Sleep Latency (min) | 25.8±14.4 | 17.2±13.8 | -1.9 (38.0) | -8.57 (4.5) | -17.6 – 0.5 | 0.063 |
| Sleep Efficiency (%) | 83.4±5.3 | 85.6±4.4 | 1.6 (38.0) | 2.5 (1.5) | -0.7 – 5.6 | 0.119 |
| WASO (minutes) | 45.7±19.1 | 35.7±12.4 | -1.9 (38.0) | -9.9 (5.1) | -20.3 – 0.4 | 0.059 |
| Sleep Duration (minutes) | 443.5±54.1 | 421.1±48.1 | -1.4 (38.0) | -22.4 (16.2) | -55.2- 10.3 | 0.174 |

**Abbreviations:** PedMIDAS, Pediatric Migraine Disability Assessment Scale; FDI, Functional Disability Inventory; PROMIS, Patient-Reported Outcomes Measurement Information System; ASWS, Adolescent Sleep Wake Scale; ASHS, Adolescent Sleep Hygiene Scale; ISI, Insomnia Severity Index; WASO, time scored as wake after sleep onset, in minutes.

**Figure 1.**
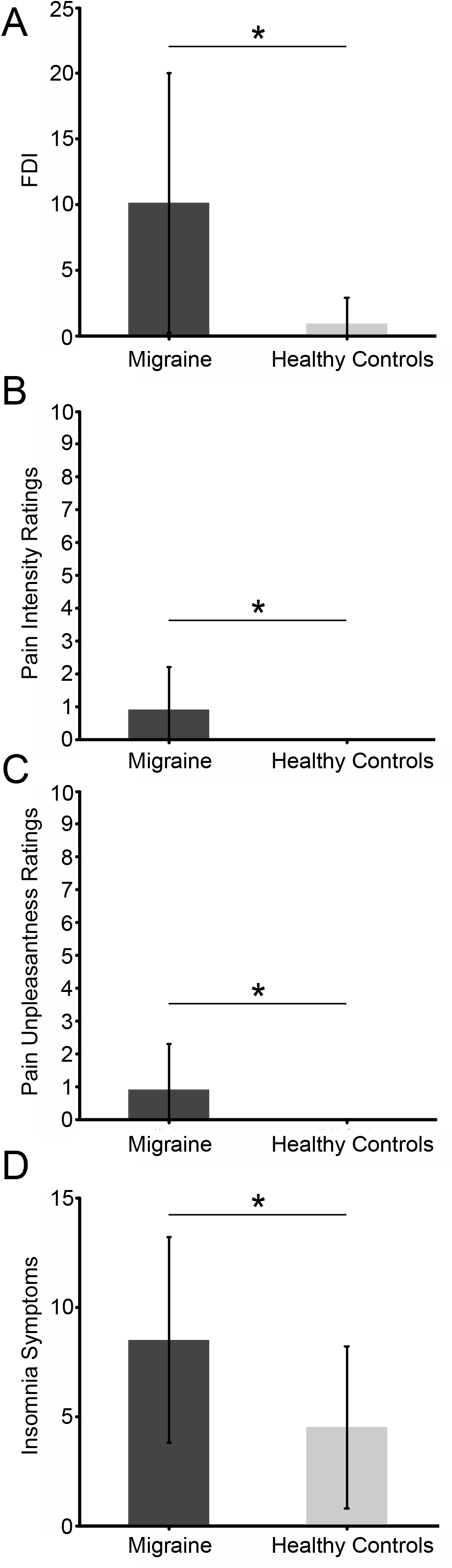
Differences between adolescents with and without migraine. Adolescents with migraine had greater FDI scores (p<0.001), greater pain intensity ratings during the scan (p=0.03), greater pain unpleasantness ratings during the scan (p=0.03), and higher insomnia symptoms (p=0.005) compared to healthy controls. FDI: Functional Disability Inventory

### Differences between adolescents with migraine and healthy controls in perceived and actigraphy sleep

Participants with migraine reported greater ISI scores (indicating greater sleep difficulties) compared to healthy controls (p=0.005, Fig. 1D). Albeit not reaching statistical significance, adolescents with migrane also showed longer actigraphy-measured SOL (+8.6 min; p=0.063) and greater amount of WASO (+10 min; p=0.059) compared to controls (Table 1). No significant group differences were found for sleep hygiene (Table 1).

Exploratory analyses examined the correlations between sleep measures and migraine frequency and disability in the migraine group. For the subjective measure of sleep difficulties, higher ISI scores (*r*=0.657, *p*=0.002) were related with greater disability (FDI) but not migraine frequency (*r*=0.419, *p*=0.066) nor PedMIDAS (*r*=0.248, *p*=0.110). No relationships with objective (actigraphic) sleep outcomes (all *p*’s>0.05) were found.

### Differences between adolescents with migraine and healthy controls in amygdalar connectivity

Adolescents with migraine, compared to healthy adolescents, had greater resting-state functional connectivity between the *left* amygdala and the bilateral posterior cingulate cortex (PCC), precuneus, secondary somatosensory cortex, primary motor cortex, premotor cortex, inferior parietal lobule, cerebellum, occipital cortex, and the left SI (Fig. 2, Table 2). For the brain areas of the posterior cingulate cortex and precuneus, no connectivity with the amygdala was found in healthy adolescents but it was found in participants with migraine. For all other regions, connectivity was found for both adolescents with migraine and healthy adolescents with stronger connectivity in participants with migraine.

**Table 2.**
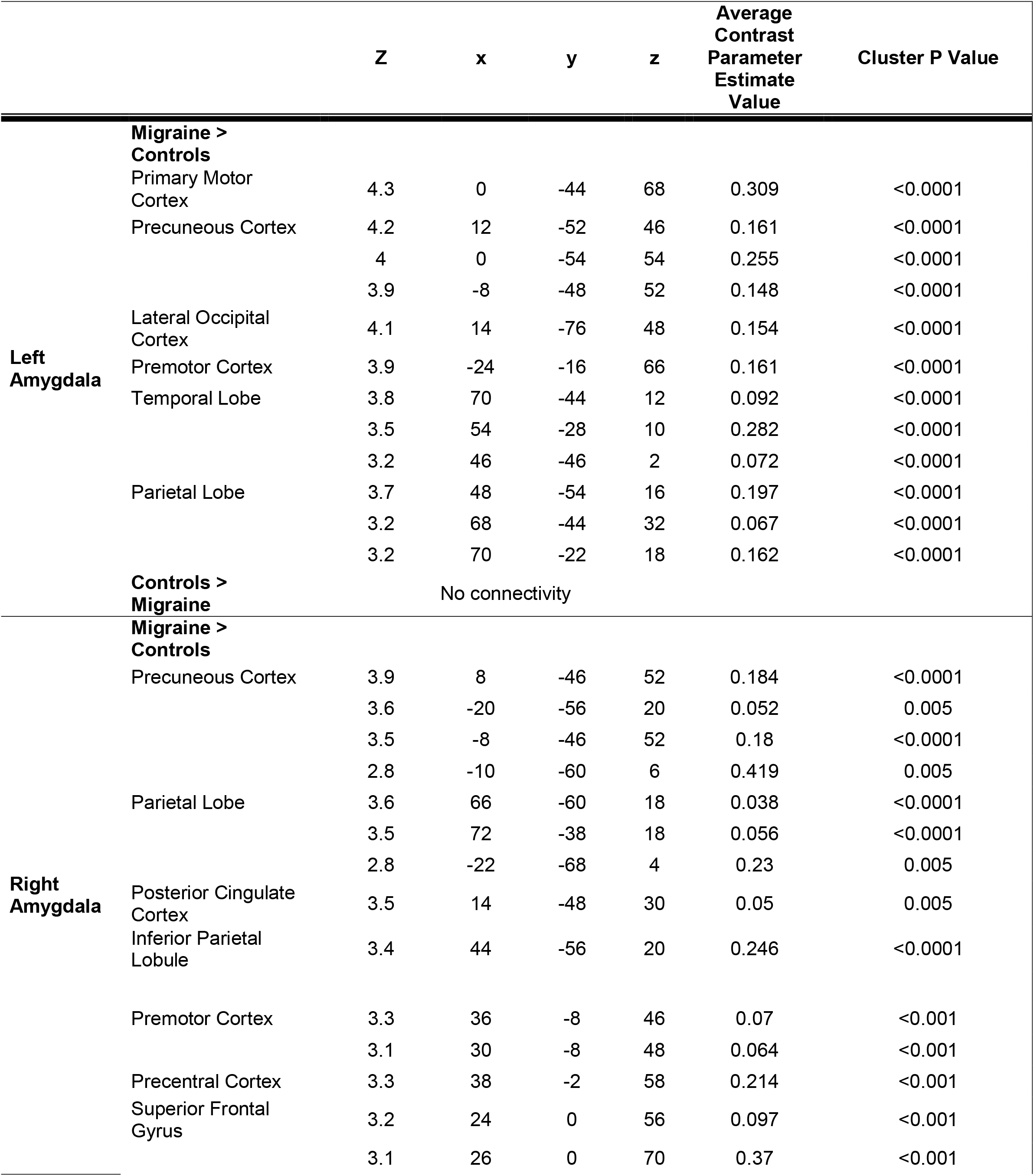
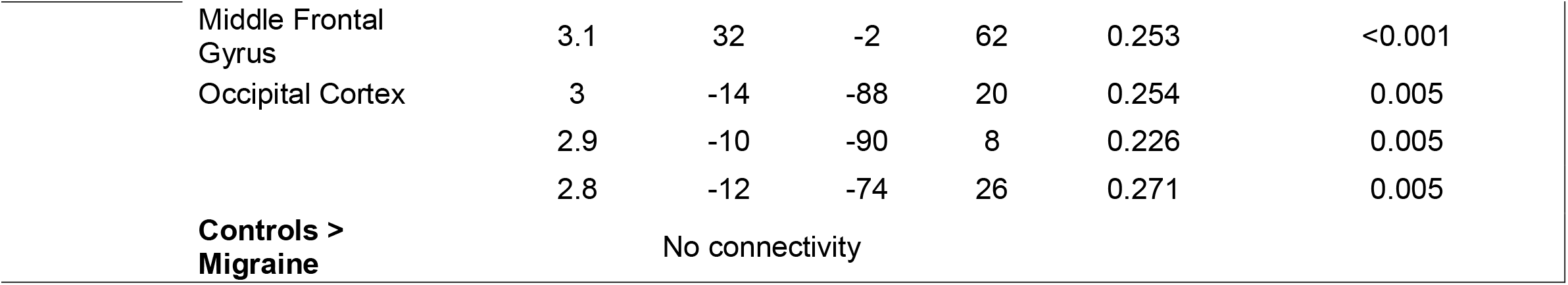
Functional Connectivity of the Amygdala in adolescents with migraine vs. healthy adolescents.

**Figure 2.**
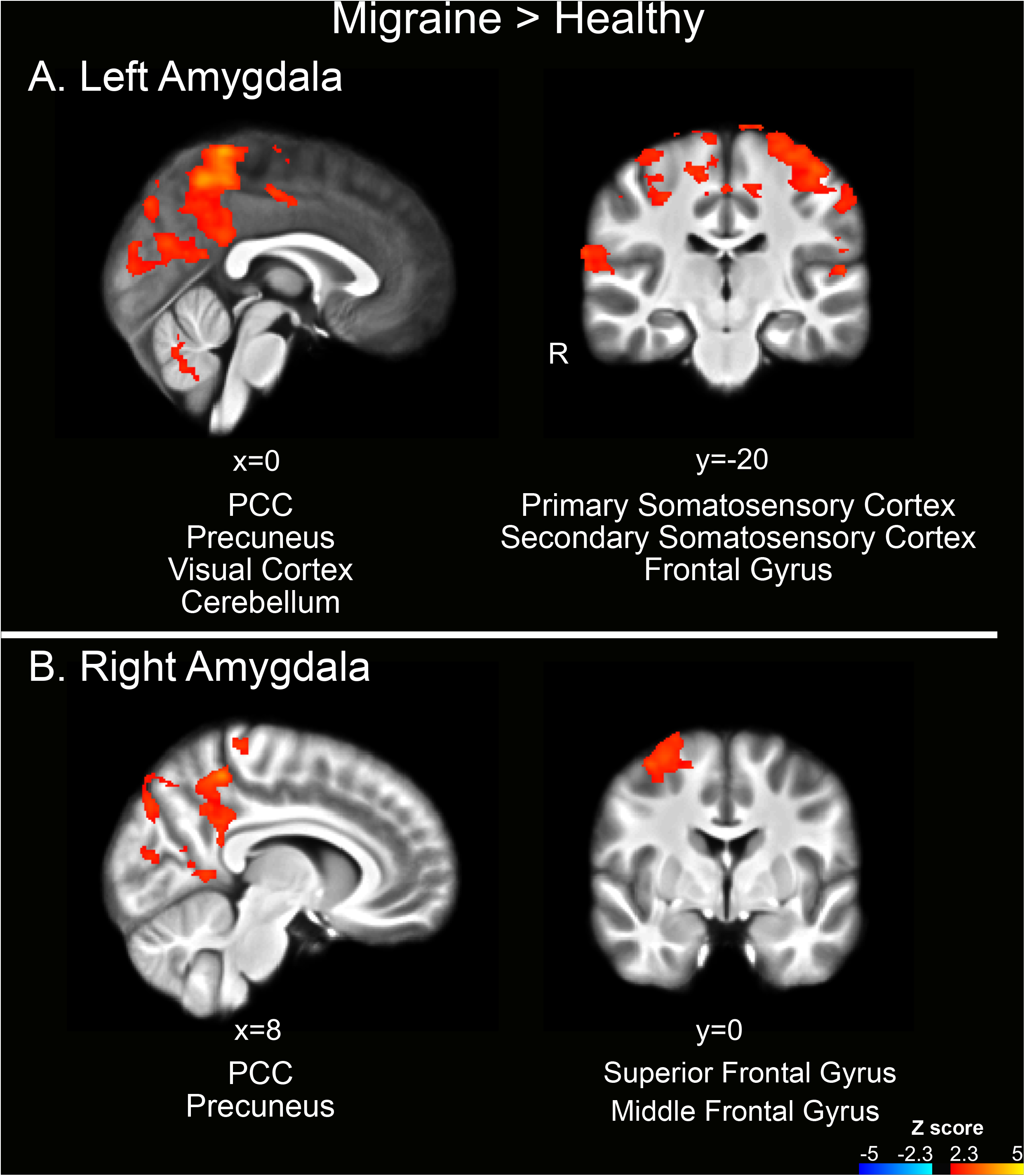
Amygdalar connectivity in adolescents with adolescents with migraine patients vs. healthy adolescents. <u>Left amygdala</u>- Greater resting-state functional connectivity was found in adolescents with migraine compared to healthy controls between the left amygdala and the bilateral PCC, precuneus, secondary somatosensory cortex, primary motor cortex, premotor cortex, inferior parietal lobule, cerebellum, occipital cortex, and the left SI. <u>Right amygdala</u>- Greater resting-state functional connectivity was found in adolescents with migraine compared to healthy controls between the right amygdala and the right PCC, superior and middle frontal gyrus, primary motor cortex, primary somatosensory cortex, inferior parietal lobule and lateral occipital cortex, as well as the bilateral precuneus and visual cortex. PCC-posterior cingulate cortex; DLPFC- dorsolateral prefrontal cortex; SI- somatosensory cortex; SII- secondary somatosensory cortex.

In addition, adolescents with migraine, compared to healthy adolescents, had greater resting-state functional connectivity between the *right* amygdala and the right PCC, superior and middle frontal gyrus, primary motor cortex, primary somatosensory cortex, inferior parietal lobule and lateral occipital cortex, as well as the bilateral precuneus and visual cortex (Fig 2). For the posterior cingulate cortex and precuneus, connectivity was found in participants with migraine but not in healthy adolescents, while for the frontal gyrus, primary motor cortex, inferior parietal lobule and lateral occipital cortex connectivity was found for both adolescents with migraine and healthy adolescents, with stronger connectivity in participants with migraine.

### Measures of perceived sleep difficulties or objective sleep disruption do not mediate the differences in amygdala connectivity between adolescents with and without migraine

Next, we tested if differences in amygdalar connectivity are mediated by the differences in sleep difficulties (Fig. 3). The difference in functional connectivity of the *left* amygdalar was not mediated by subjective sleep difficulties (ISI, estimate 0.004, 95 % CI [-0.029] – [0.024]) or objective actigraphic sleep disruption (WASO, estimate -0.001, 95 % CI [-0.025] – [0.024]). Similarly, the difference in functional connectivity of the *right* amygdalar was not mediated by ISI (estimate 0.013, 95 % CI [-0.020] – [0.037]) or WASO (estimate -0.008, 95 % CI [-0.036] – [0.016]).

**Figure 3.**
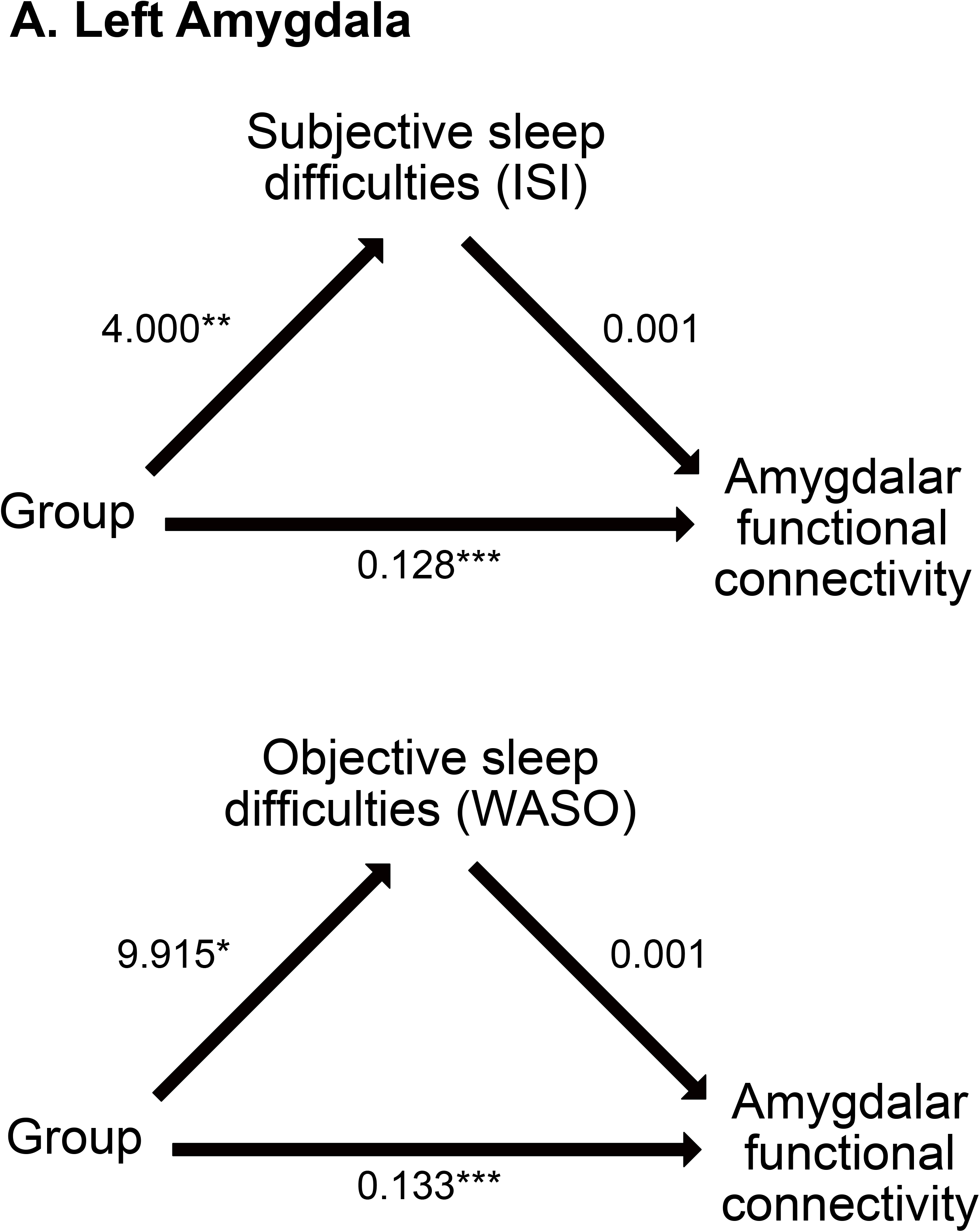
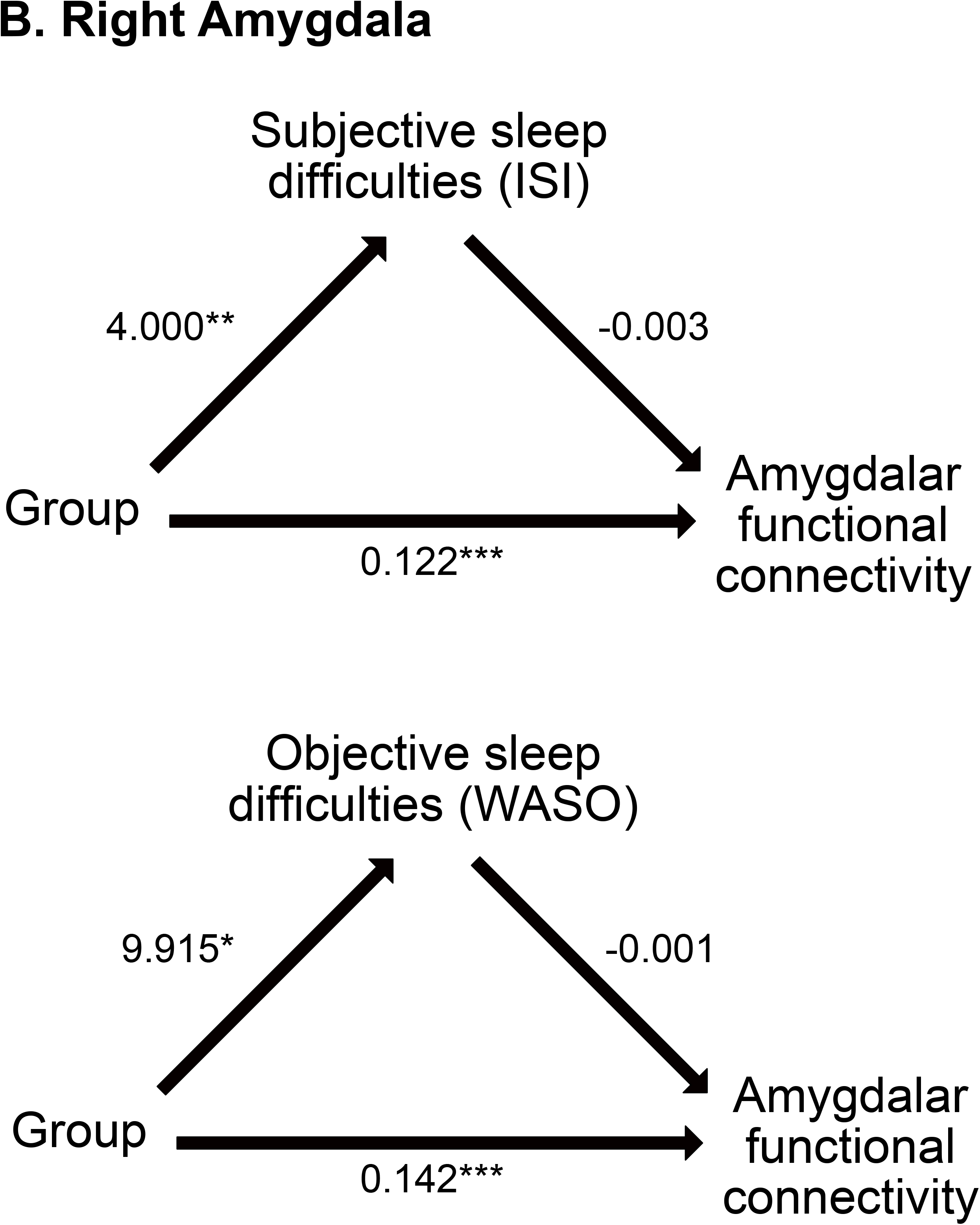
Subjective and objective sleep difficulties do not mediate the group differences in amygdalar connectivity. Group differences were found for functional connectivity of the amygdala and sleep difficulties. However, sleep difficulties were not related to functional connectivity of the amygdala (left (**A**) or right (**B**). The numbers for each path represent the unstandardized estimates for each path in the mediation model. Each mediation model was analyzed separately. * p<0.05, ** p<0.01, ***p<0.001. ISI: Insomnia Severity Index; WASO: wake minutes after sleep onset (time scored as wake after sleep onset, in minutes)

## Discussion

This study found differences in sleep difficulties and alterations in amygdalar connectivity between adolescents with and without migraine. Compared to healthy adolescents, adolescents with migraine exhibited greater sleep difficulties and showed greater connectivity between the amygdala and somatosensory and frontal areas which are involved in sensory and cognitive aspects of pain. However, measures of perceived sleep difficulties or actigraphic sleep disruption did not mediate the differences between the groups in amygdalar connectivity.

### Differences in sleep measures

This study extends previous research regarding sleep disturbances in children and adolescents with chronic pain conditions (55, 56) including migraine (23, 24). Due to the multidimensionality of sleep, we assessed a number of domains previously shown to be impaired in pediatric pain patients. In our current cohort, adolescents with migraine compared to controls spent similar amount of time asleep and did not significantly differ in sleep habits (sleep hygiene). However, perceived sleep difficulties (i.e. insomnia symptoms including difficulty falling asleep and maintaining sleep) were greater in those adolescents with migraine. Similarly, actigraphic sleep assessment showed a tendency for greater sleep disruption (amount of WASO) in adolescents with migraine.

It is worth mentioning that in the current sample, the perceived sleep difficulties (reflecting insomnia symptoms) of adolescents with migraine, may not necessary reaching clinical significance. When considering the ISI cut offs for subthreshold and clinical insomnia (adults (40)), the majority of adolescents with migraine (n = 10, 50%) would be considered to have no clinically significant insomnia (ISI, < 8). However, no clinical cutpoints for ISI have been validated in adolescents, and a standardized clinical assessment of insomnia (57) would be ultimately necessary to clinically determine the extent of the sleep difficulties reported by adolescents with migrane. Thus, it is possible that differences in connectivity could be more prominent in the presence of adolescence with migrane and clinical significant insomnia.

Insomnia is the most common sleep disorder in adolescents (57); it is common in pediatric pain leading to poorer health-related quality of life, greater disability and greater psychological distress (18, 37, 58-60). Despite the etiology of the disorder is not fully understood, insomnia is characterized as an hyperarousal disorder in which several psychophysiological domains are hyperactivated and are thought to interfere with the falling asleep and sleep processes of insomniacs (57). It is possible that the presence of insomnia leads to a greater tendency for greater hyperarousal (61) (62) (60, 63) before and during the sleep period, which results in reductions in emotional functioning and consequently may increases the risk for elicitation, persistence, and severity of headaches in adolescents with migraine. Furthermore, treating insomnia has been shown to improve outcomes in adolescents with co-occurring pain and insomnia (64). A recent study demonstrated the impact of cognitive-behavioral therapy for insomnia in adolescents with migraine – showing a reduction in headache days (commonly used to guide and track clinical treatment) and insomnia symptoms (65). Overall, the presence of insomnia in adolescents with migraine may offer a key target for intervention.

### Differences in functional connectivity between adolescents with migraine vs. healthy controls

Increases in amygdalar connectivity with the SI, PCC, and precuneus were found in adolescents with migraine compared to healthy controls. The amygdala is involved in nociceptive processing (66, 67) but also in pain attention and anticipation (68) (69). Amygdalar activation can be reduced when participants shift their attention by performing a task while waiting for a noxious stimulus (68). Similarly, an increased response in the amygdala has been found during attention to the unpleasantness of a noxious stimulus (70). Another brain region that is involved in pain anticipation is the precuneus (71). Thus, the greater functional connectivity between the amygdala and the precuneus may represent altered affective-cognitive functions of pain attention and anticipation in adolescents with migraine. Both children and adults with migraine, perform worse on cognitive tests including attention tasks (72) (73-76). On the other hand, patients with migraine have enhanced response to somatosensory attention tasks. Adults with migraine had similar laser evoked potentials with and without a distraction arithmetic task while in healthy controls, a reduction in laser evoked potentials was found during the task (77). Similarly, in adolescents with migraine, somatosensory stimuli (both noxious and non-noxious) that were delivered with auditory stimuli (target for attention) yielded larger p300 amplitudes (78). Thus, patients with migraine might have a greater effect of the amygdala on areas involved in affective and cognitive regulation of pain. This could result or contribute to the attentional bias of individuals with migraine towards somatosensory stimuli (both noxious and non-noxious) (78) and to the constant worry and anticipation for the next headache attack (79).

Only a few studies have examined neural alterations between youth with migraine and healthy controls. Faria and colleagues found that female adolescents with migraine had greater connectivity between the left amygdala and the right supplementary motor area, right precuneus, bilateral thalamus, bilateral anterior cingulate cortex and left insula compared with males adolescents with migraine and healthy controls (7). In another study from the same group, the resting-state functional connectivity of brain networks was examined. Different patterns of connectivity were found between youth with migraine and healthy controls in various networks including the central executive, salience, default mode, auditory, frontal-parietal and sensorimotor networks (9). In the same cohort but using a different method of cerebral blood flow, alterations in regional cerebral blood flow during resting-state were examined in pediatric and young adult participants with migraine and healthy controls (age range 8-24). Greater cerebral blood flow in the bilateral primary somatosensory cortex was found in those with migraine compared with healthy controls (8). Overall, these results are not in line with the results of the present study; comparison between the studies is difficult due to the differences in analysis methods (functional connectivity vs. activation; seed-based connectivity vs. network connectivity). Nevertheless, our results are similar to the results of other studies conducted in adults with migraine in which increased functional connectivity between the amygdala (left and right) and the secondary somatosensory cortex (80), as well as between the left amygdala and the middle cingulate and precuneus (25) and superior frontal (81) are observed in those with migraine compared to adult healthy controls.

### Measures of perceived sleep difficulties and objective sleep disruption do not mediate the differences in amygdalar connectivity between the groups

The groups differed in both perceived sleep difficulties and resting state functional connectivity of the amygdala, while also showing a tendency for having greater objective sleep disruption. While involved pain, the amygdala also has a role in sleep processes. Animal studies have found that the central nucleus of the amygdala has glutamatergic projections to neurons in the nucleus pontis oralis that may generate active sleep (82, 83). In humans, after 36 hours of total sleep deprivation, increased connectivity between the amygdala and the PCC/precuneus was found (84), and alterations in amygdalar connectivity and activation are found in people with insomnia (85-88). In addition, adults with migraine had different relationships between amygdalar connectivity and sleep disturbance compared to healthy controls (25). Thus, since amygdalar connectivity and, specifically, its connectivity with the precuneus is related to both migraine and sleep disorders, it may be involved in the effects of sleep on pain or, vice versa of pain on sleep.

However, even though sleep dificulties were greater in adolescents with migraine, sleep dificulties did not mediate the differences in functional connectivity between adolescents with migraine and healthy adolescents. This was found for both subjective (ISI) and objective (WASO) measures for sleep difficulties and disruption, respectively. Thus, it is suggested that the changes between the groups in amygdalar connectivity are not due to differences in perceived sleep difficulties or actigraphic sleep disruption. To mention, these results need to be contextualized in light of the limitations of actigraphy. Actigraphy allows to discriminating sleep and wake based on the individuals’ pattern of motion (motion = wake; absence of motion = asleep), and not capturing the full expression of sleep (e.g., sleep stages, arousals). In addition, actigraphic wake has been frequently underestimated by actigraphy, mainly due to the failure of actigraphy in discriminating motionless wake from sleep (89). Future studies based on gold standard laboratory sleep assessments (including measures of polysomnographic sleep quality and continuity, and electroencephalographic measures of sleep quantification) may further elucidate the relation migrane – sleep – functional connectivity.

## Conclusions

The observed alterations in amygdalar connectivity with areas involved in cognitive function and nociceptive processing and the alterations in sleep may play a role in migraine pathophysiology in pediatric patients. However, these alterations may be independent.

## Acknowledgments

We thank Thomas Maloney (Cincinnati Children’s Hospital) for his aCompCor scripts. We thank Dr. Marielle Kabbouche Samaha, Dr. Hope L. O’Brien, Dr. Joanne Kacperski, Jessica L. Weberding, Susan L. LeCates, Mimi N. Miller and Shannon Kathleen K. White of the Cincinnati Children’s Headache Center. We thank Dr. Tonya Palermo for providing the sleep diary and guidance for the sleep actigraphy. The study was funded by the Cincinnati Children’s Hospital Discovery Award and startup funds for RCC and CDK.

